# Systematic review automation tool use by systematic reviewers, health technology assessors and clinical guideline developers: tools used, abandoned, and desired

**DOI:** 10.1101/2021.04.26.21255833

**Authors:** Anna Mae Scott, Connor Forbes, Justin Clark, Matt Carter, Paul Glasziou, Zachary Munn

## Abstract

**Objective:** We investigated the use of systematic review automation tools by systematic reviewers, health technology assessors and clinical guideline developers.

**Study design and settings:** An online, 16-question survey was distributed across several evidence synthesis, health technology assessment and guideline development organisations internationally. We asked the respondents what tools they use and abandon, how often and when they use the tools, their perceived time savings and accuracy, and desired new tools. Descriptive statistics were used to report the results.

**Results:** 253 respondents completed the survey; 89% have used systematic review automation tools – most frequently whilst screening (79%). Respondents’ ‘top 3’ tools include: Covidence (45%), RevMan (35%), Rayyan and GRADEPro (both 22%); most commonly abandoned were Rayyan (19%), Covidence (15%), DistillerSR (14%) and RevMan (13%). Majority thought tools saved time (80%) and increased accuracy (54%). Respondents taught themselves to how to use the tools (72%), and were most often prevented by lack of knowledge from their adoption (51%). Most new tool development was suggested for the searching and data extraction stages.

**Conclusion:** Automation tools are likely to take on an increasingly important role in high quality and timely reviews. Further work is required in training and dissemination of automation tools and ensuring they meet the desirable features of those conducting systematic reviews.

## Introduction

Systematic reviews are integral to evidence-based decision making. They serve as the input into clinical and policy decision-making, both on their own and as underpinnings of clinical practice guidelines and health technology assessments. However, conducting them has historically been both resource- and time-intensive, with systematic reviews on average, requiring approximately 67 weeks to complete,(1) guidelines requiring 18-30 months,(2) and health technology assessments between 6 and 24 months(3).

To decrease the time required, several automation tools have been – and continue to be – developed, to assist with completing the key systematic review stages, such as devising and conducting database searches, screening of search results, data extraction, meta-analysis, and write up of the results.(4-9)

Although automation tools are potentially helpful, their current uptake appears low. However, this evidence based on a few subgroups of the systematic reviewer community, for example, Cochrane systematic reviewers,(10) and clinical practice guideline developers who are members of Guideline International Network.(11)

To broaden our understanding of the uptake of automation tools, we conducted a survey to identify whether there are differences in how those who conduct standalone systematic reviews, clinical practice guidelines, and health technology assessments (henceforth, collectively ‘reviews’) perceive and interact with systematic review automation tools. More specifically, we queried what types of tools they use and have abandoned, how often they use the tools, at what stages of the process, how they perceive the time savings and accuracy of the tools, how they learn to use the tools, and what new tools they would like developed.

## Methods

We conducted a survey of self-identified systematic reviewers, health technology assessors and guideline developers, assessing review experience and views and experiences with the use of automation tools using a set of multiple choice and open-ended questions. This survey is reported following the Checklist for Reporting Results of Internet E-Surveys (CHERRIES) reporting guideline.(12)

### Respondents

The survey targeted respondents engaged in conducting any part of systematic reviews, clinical practice guidelines, and health technology assessments. The survey was “open,” i.e., anyone could complete it. We did not impose any location, gender, or age restrictions on the respondents, although we had anticipated that respondents would be over 18, as the questions targeted professionals.

### Survey dissemination

We adopted a two-pronged approach to reach respondents: via professional organisations and via social media.

We contacted professional organisations whose membership includes desired respondents via email, with a request to disseminate it to their membership. The email provided information about the project and its aims, as well as a link to the survey. The following organisations were contacted: JBI (formerly known as the Joanna Briggs Institute, a systematic review organisation), Cochrane Collaboration (a systematic review organisation), G-I-N: Guidelines International Network (guidelines-focused), HTAi (health technology assessment international, HTA-focused), INAHTA (International Network of Agencies for Health Technology Assessment (HTA-focused), ALIA Health Libraries Australia (librarians involved in searching for systematic reviews), and Expert Searchers Mailing list (international; individuals involved in designing/executing searches for systematic reviews).

We also disseminated the information about the survey via personal Twitter accounts (AMS, PG, ZM), and an institutional account (Institute for Evidence-Based Healthcare), with weekly tweets with information about the survey and a link.

The survey was opened on 28 Sept 2020; all survey responses were eligible for inclusion if returned before 31 October 2020.

### Survey instrument

The survey was deliberately brief, consisting of 16 questions plus an option to provide an email address to receive results. The questions were predominantly tick-box or multiple-choice questions, querying the respondent’s experience with systematic reviews, usage of automation tools, perception of their accuracy and time savings offered, and how they learned to use the tools. (The full survey is reproduced in Appendix A).

To test for technical issues and estimate the time required to complete the survey, we piloted the survey with two colleagues not involved with the project. Based on the feedback received, the survey was reformatted to decrease the need for scrolling, and the resulting survey consisted of 7 pages. The estimated time for completion was approximately 10 minutes. There was only one version of the survey, and the sequence of questions was constant – i.e., adaptive questioning was not used – but respondents were able to return to prior answers to change them.

The survey was hosted on the SurveyMonkey platform.

### Analyses

Excel was used to calculate descriptive statistics. We had intended to calculate the difference in proportions between groups using the chi-square test, however, data was insufficient. Free text responses were analysed thematically, using an inductive approach.(2) Coding was conducted by two researchers (AMS, CF), with discrepancies resolved by consensus.

### Ethics approval and informed consent

Bond University Human Research Ethics Committee provided approval for the project (AS200903). The first page of the survey provided information about the project, project team, ethics approval, data protection, and consent. Respondents provided their consent by clicking on the “if you consent, please click next to proceed” button. Participation and completion were voluntary, and no incentives for survey completion were offered.

## Results

### Respondents

We received 253 responses. The median time to completion was 6 minutes. As the survey was open to all those with experience in evidence synthesis, no response rate was able to be calculated.

Respondents reported conducting reviews via their own organisation (e.g., an employer) (59%), JBI (15%), Cochrane (13%), or another affiliation (13%). Respondents who identified another affiliation and provided additional details, indicated conducting reviews for multiple organisations, government agencies, HTA agencies, guideline-producing bodies, or universities.

The majority of respondents had been involved in more than 10 reviews (53%); 13% had previously conducted between 6-10 reviews, 23% between 3-5, 9% between 1-2 reviews. 1% reported having not been involved in previous reviews.

### Types of reviews conducted

Nearly two-thirds of respondents conducted systematic reviews as stand-alone projects (66%); 21% conducted them as part of a guideline development process, and 13% as part of a health technology assessment.

The majority of respondents had conducted reviews of interventions (82%), then, in descending frequency: scoping reviews (46%), qualitative reviews (29%), diagnostic test accuracy reviews (25%) and prognostic reviews (17%). All other review types (including: aetiology, prevalence, economic, health utilities, patient preferences, and other) were conducted by fewer than 15% of respondents. (Appendix B, Table B1).

Respondents most frequently reported involvement in the search design/execution (78%) and question-formulating stages of a review (72%). However, also common were involvement in the write-up (59%), screening (54%), data extraction (51%) and data synthesis (45%) stages.

### Frequency and stage of automation tool use

202 respondents reported the percent of their reviews that involved use of automation tools; 11% of respondents reported using automation tools in none of their reviews while 89% used automation tools in at least some of their reviews in the past 3 years.

Automation tools were most frequently used at the screening stage (79% of 189 respondents), followed by data extraction (51%) and data synthesis/meta-analysis stages (46%) (Table 1).

**Table 1:**
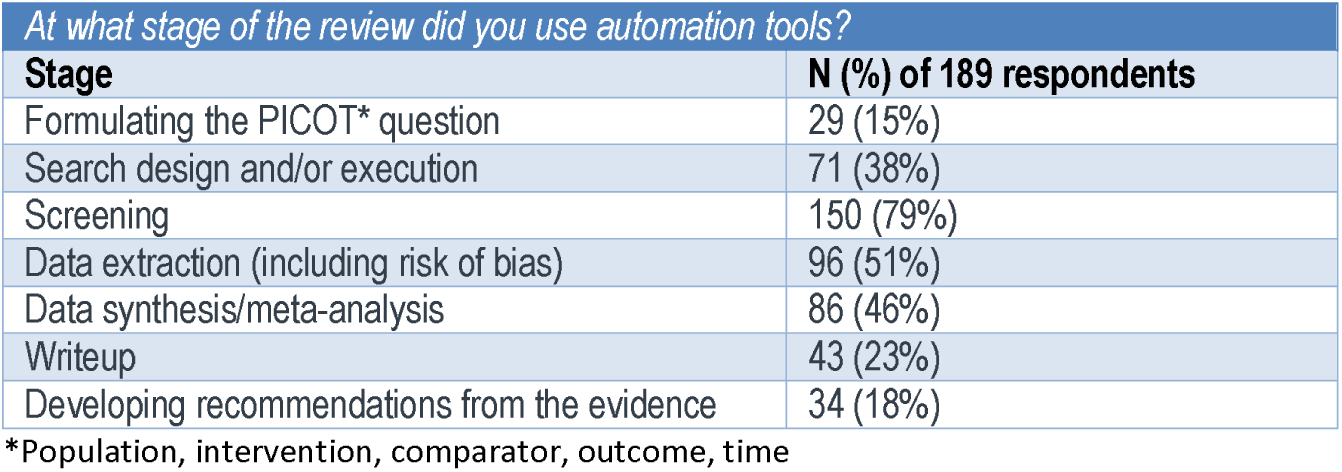
Stage of automation tool use.

### Automation tools used and abandoned

211 respondents provided information on the tools they used during the systematic review process. Most commonly selected tools from a list of 45 options (including ‘other’ with an option to provide details), included: Covidence (50%), RevMan (41%), Rayyan (33%), GRADEpro (31%) and JBI-SUMARI (21%) (Figure 1; Figure B1 in Appendix B).

**Figure 1:**
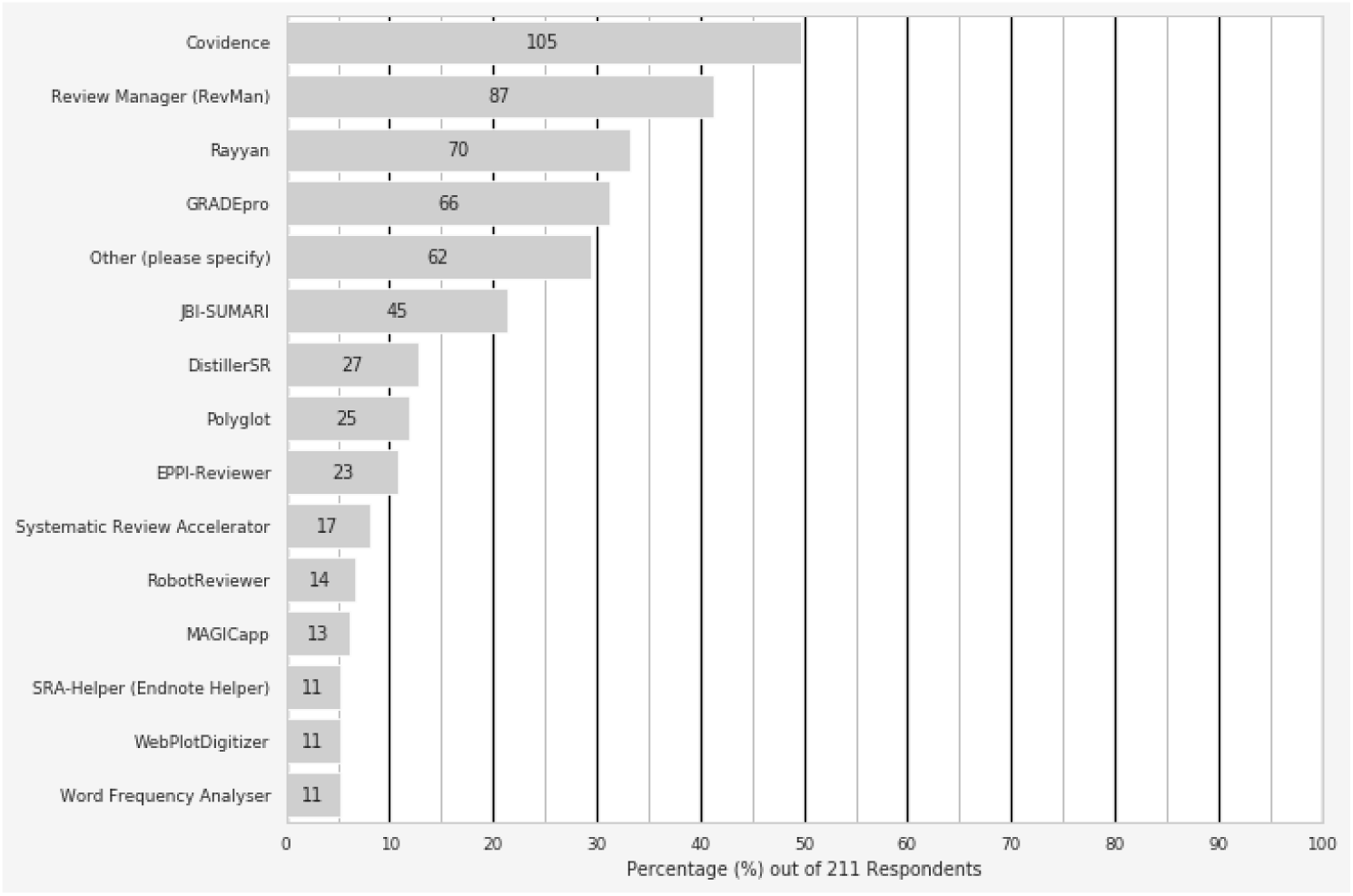
Fifteen most commonly used systematic review automation tools.

Respondents were also asked to identify their ‘top 3’ – the three tools they use most commonly. Those used most commonly by 180 respondents, included: Covidence (45%), RevMan (35%), and Rayyan and GRADEPro (both 22%).

The top tools reported to be abandoned most frequently by 95 respondents included Rayyan (19%), Covidence (15%), DistillerSR (14%) and RevMan (13%). Reasons for abandoning the use of these tools included: crashes, non-customisability, slowness, cost, complexity or difficulty to learn, preference for or availability of an alternate tool, and lack of desired features.

### Speed and accuracy improvements

Eighty percent (80%) of 184 respondents thought that automation tools save time – either a lot of time (36%) or some time (44%); 15% were neutral and 4% thought there are time costs associated with tool use (Table 2). Most frequently, respondents were neutral about the improvement in accuracy from automation tool use (44% of 185 respondents), although 54% felt there was either a lot or a little accuracy improvement. 2% thought there is some accuracy loss from the use of automation tools (Table 2).

**Table 2:**
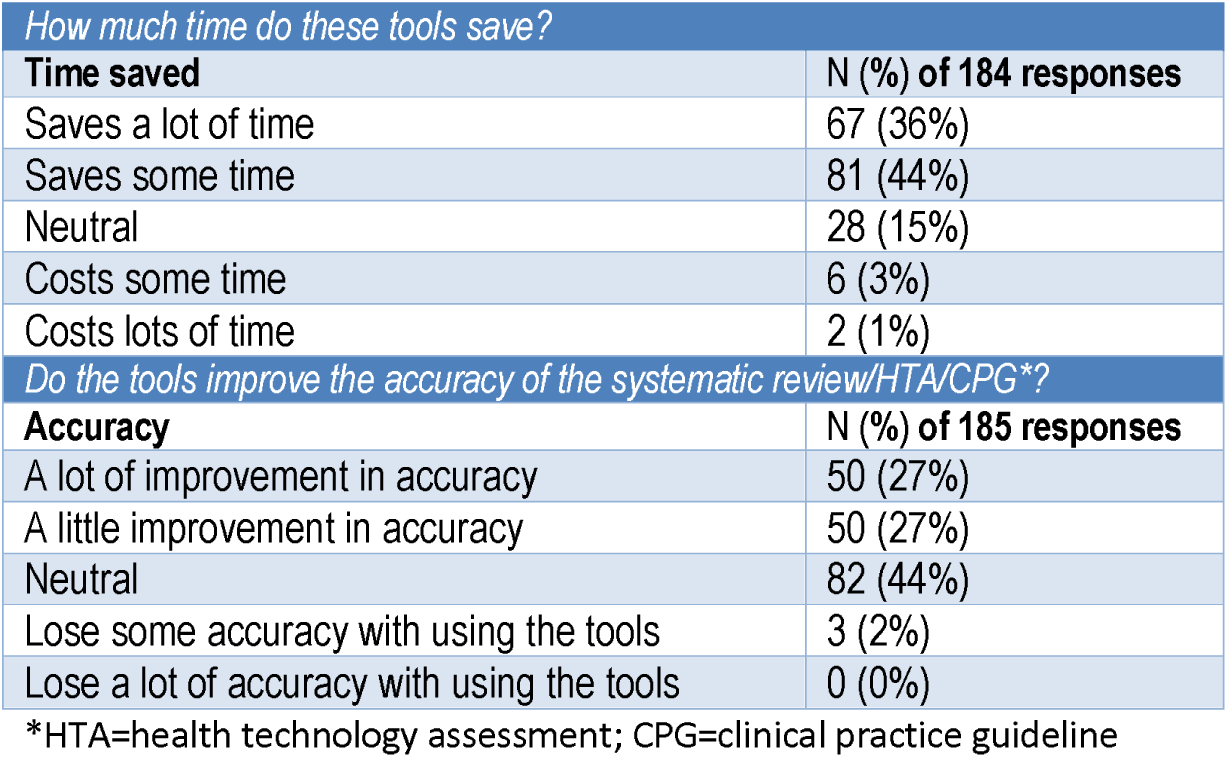
Speed and accuracy improvement from automation tool use.

| *How much time do these tools save?* |  |
| --- | --- |
| **Time saved** | **N (%) of 184 responses** |
| Saves a lot of time | 67 (36%) |
| Saves some time | 81 (44%) |
| Neutral | 28 (15%) |
| Costs some time | 6 (3%) |
| Costs lots of time | 2 (1%) |
| *Do the tools improve the accuracy of the systematic review/HTA/CPG\*?* |  |
| **Accuracy** | **N (%) of 185 responses** |
| A lot of improvement in accuracy | 50 (27%) |
| A little improvement in accuracy | 50 (27%) |
| Neutral | 82 (44%) |
| Lose some accuracy with using the tools | 3 (2%) |
| Lose a lot of accuracy with using the tools | 0 (0%) |

\*HTA=health technology assessment; CPG=clinical practice guideline

### Learning about the tools and factors impeding their use

Most commonly, respondents taught themselves how to use the automation tools (72% of 193 respondents) or used the help documentation (44%). Learning from others commonly involved participating in workshops (43%), learning from a colleague (42%) or from webinars (40%) (Figure 2).

**Figure 2:**
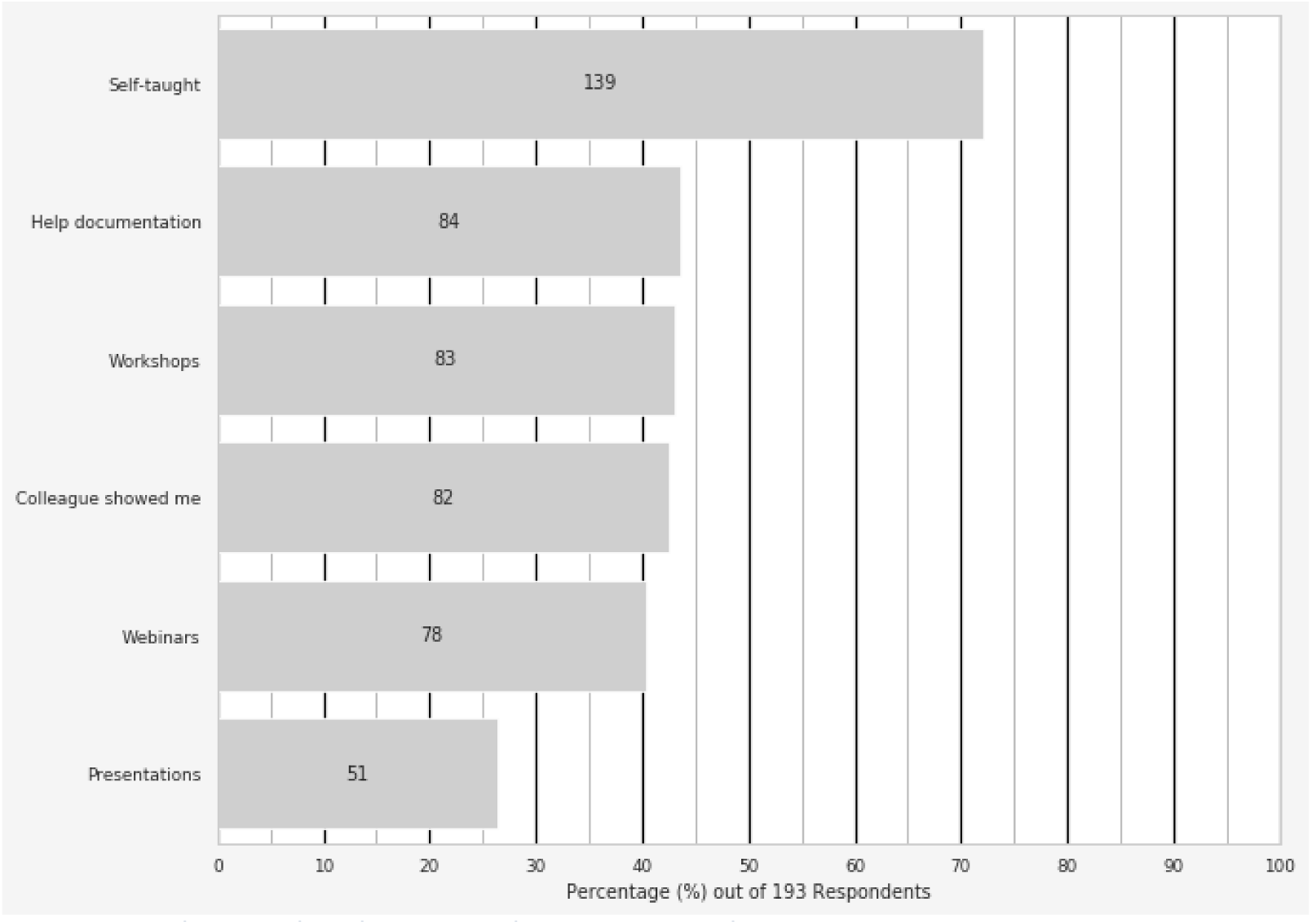
How do respondents learn to use the automation tools.

Respondents (n=196) were most commonly prevented from using automation tools by lack of knowledge about the existing tools (51%), costs (45%), the complicated nature of the tools (39%), time constraints (28%), or user unfriendliness (25%). (Table 3)

**Table 3:** Factors preventing respondents from using the automation tools.

| What prevents respondents from using the automation tools |  |
| --- | --- |
| Factors | N (%) of 196 responses |
| The learning curve (too complicated to learn) | 76 (39%) |
| Time constraints | 55 (28%) |
| Costs | 88 (45%) |
| Language barrier | 5 (3%) |
| Internet connectivity/firewall | 5 (3%) |
| User unfriendly | 49 (25%) |
| Lack of knowledge of existing tools | 100 (51%) |
| Not applicable | 20 (10%) |
| Other (optional: please share why) | 43 (22%) |

### Most time-consuming steps and new tools desired

Of 194 respondents, 48% found screening to be the most time-consuming stage of the review, followed by data extraction (45%) and search design and/or execution (30%). Data synthesis/meta-analysis was identified by 19% of respondents, and write-up by 16%; fewest respondents found formulating the PICO question to be the most time-consuming step (8%).

Respondents also shared what automation tools they would like to see developed in the future. As multiple suggestions were frequently made in a single comment, 116 comments were broken down to individual suggestions (n=341). Most commonly, tools for the searching stage (53 suggestions), data extraction stage (n=41), screening (n=20) or risk of bias (n=19) were requested. (The specific tools requested by more than one respondent are in Table 4; an expanded table is presented in Appendix B, Table B2).

**Table 4:** Tool development suggested by the respondents.

| *What automation tools would you like developed?* |  |
| --- | --- |
| **Review stage (N suggestions)** | **Suggested tools\*** |
| Searching (n=53) | • Strategy optimisation tool (non-specific or visual) (n=14)<br>• Deduplication (n=9)<br>• Translation between databases (n=7)<br>• Strategy design (non-specific or data mining tools) (n=5)<br>• Grey literature (non-specific or exporting) (n=2)<br>• Synonym/related term (phrase) suggestion tool (n=2) |
| Screening (n=20) | • Dispute resolution tool (non-specific or with visualisation) (n=5)<br>• Screening tool (non-specific) (n=4)<br>• Data mining functionality (for full-text screen) (n=2)<br>• Reference prioritisation and threshold-setting functionality (n=2) |
| Data extraction (other than risk of bias) (n=41) | • Data extraction tool (non-specific or general - e.g., accurate, fast) (n=24)<br>• For observational studies (n=2)<br>• For reuse of previous data extractions (n=2)<br>• From Pdfs (n=2) |
| Risk of Bias (n=19) | • Automated risk of bias (non-specific) (n=11)<br>• Automated risk of bias (observational studies) (n=7) |
| Synthesis/analysis (n=5) | • No tools suggested by more than 1 respondent |
| Writeup (n=12) | • Data visualisation/presentation (n=5) |
| Project management (n=16) | • Streamlined/single tool for the entire process (n=9)<br>• Linkage between tools/data pushing between tools (n=2) |
| Other (n=4) | • No tools suggested by more than 1 respondent |

\*the number of suggestions for each tool is indicated in brackets; complete list of suggested tools is provided in Table B2, Appendix B

### Difference in responses by systematic reviewers, guideline developers and health technology assessors

Paucity of responses from guideline developers and health technology assessors precluded testing whether the three categories of respondents differ in their perception of the usefulness of the automation tools. We therefore present the percentages of responses in each category.

Most systematic reviewers saw automation tools as offering time savings (80%) – either some time (43%) or a lot of time (37%); very few saw automation tools as involving time costs (3%). Similarly, the majority of guideline developers (88%) and health technology assessors (73%) saw automation tools as offering some or large time-savings, and few perceived time costs (6% and 7%, respectively). However, the low numbers of responders in the latter categories suggest caution in interpreting these numbers. (Table 5).

**Table 5:** Perception of time-savings offered by automation tools by the 3 respondent groups.

| Time savings by automation tools | Systematic reviewers (stand-alone) N (%) of 136 responses | Guideline developers N (%) of 32 responses | Health technology assessors N (%) of 15 responses |
| --- | --- | --- | --- |
| **Costs lots of time** | 2 (1%) | 0 (0%) | 0 (0%) |
| **Costs some time** | 3 (2%) | 2 (6%) | 1 (7%) |
| **Neutral** | 23 (17%) | 2 (6%) | 3 (20%) |
| **Saves some time** | 58 (43%) | 14 (44%) | 8 (53%) |
| **Saves lots of time** | 50 (37%) | 14 (44%) | 3 (20%) |

Most systematic reviewers thought that automation tools improve accuracy (53%) – either ‘some’ (28%) or a lot (25%), however a considerable percentage were neutral (46%). A similar response pattern was evident for guideline developers, 45% of whom saw automation tools as offering a lot of improvement and 23% as offering some improvement in accuracy, whilst 32% were neutral. Health technology assessors were most commonly neutral about the improvement offered by the tools (53%) although 27% viewed them as offering some and 13% offering a lot of improvement in accuracy. The low number of responders who self-identified as guideline developers or health technology assessor dictate caution in interpretation of these numbers, however.

| Accuracy improvement by automation tools | Systematic reviewers (stand-alone) N (%) of 138 responses | Guideline developers N (%) of 31 responses | Health technology assessors N (%) of 15 responses |
| --- | --- | --- | --- |
| **A lot of loss of accuracy** | 0 (0%) | 0 (0%) | 0 (0%) |
| **Some loss of accuracy** | 2 (1%) | 0 (0%) | 1 (7%) |
| **Neutral** | 63 (46%) | 10 (32%) | 8 (53%) |
| **Some improvement** | 39 (28%) | 7 (23%) | 4 (27%) |
| **A lot of improvement** | 34 (25%) | 14 (45%) | 2 (13%) |

## Discussion

This survey of systematic reviewers, clinical practice guideline developers and health technology assessors found that most use only a few tools but consider that they save them time, though they also often abandoned use of the tools. Most commonly, respondents self-taught to use the tools, and the biggest barrier to uptake was the lack of knowledge about the tools’ existence, identified by half of the respondents.

Covidence, Rayyan and RevMan were identified as both most commonly used – and most commonly abandoned – tools. The pervasiveness of their use may be explained by their cross-utilisation and name-recognition (e.g., RevMan, and Covidence are standardly used for Cochrane Reviews, with Covidence also having partnered with JBI and the Guidelines International Network). The reasons for these tools’ abandonment by the users – technical, non-customisability, slowness, cost, and complexity – are generally consistent with the more general barriers to automation tool use identified by respondents – cost, complexity, time constraints, user-unfriendliness and technical issues. Respondents also reported high level of involvement across all of the stages of the review process (with the lowest percentage – just under a half – involved in data synthesis), however, the most common use of automation tools by far (80% of respondents) was reported in the screening stage. This may reflect the availability with the tools for this stage of the process,(13) and is reflected in the suggestions for tools to be developed – with most focusing on the data extraction or searching stages, and less commonly on screening. Interestingly, several of the tools suggested for development by the respondents already exist, which further underscores the importance of another of the survey’s finding – that lack of knowledge about the tools’ existence is the most frequently cited barrier to their adoption.

The survey has several limitations. First, one of the key limitations was the relatively low number of responses from guideline developers (21%) and health technology assessors (13%), which precluded us from identifying the differences (if any) between these three groups in their views and practices around automation tools. Second, the respondent sample may be biased and thus limited in generalisability, as those individuals who responded to our survey may generally be more interested and thus more positively inclined towards automation tool use, than non-respondents. Finally, because the survey was conducted and publicised in English, its findings may also not be generalisable to reviewers who work in other languages, although some of the ‘other’ responses to the question about the organisation through which they perform reviews, suggest that non-English-language based respondents also participated in the survey (including from: Spain, Quebec, Argentina, Norway, Austria, Croatia, Basque region, and Switzerland).

Our findings are consistent with previous findings. A recent survey focused on the automation tool use by guideline developers specifically, found that 74% use automation tools to be more efficient and 66% strongly agreed that automation tools are useful.(11) Similarly, a survey of systematic reviewers who conduct Cochrane Reviews found a usability score greater than 68 (out of 100) across automation tools.(10) The specific tools cited as most commonly used by the respondents to the present survey – Covidence, RevMan and Rayyan – also overlap with those identified by guideline developers (RevMan, Covidence and Rayyan)(11) and those identified by Cochrane Reviewers (EndNote, RevMan, Covidence, Rayyan).(10) A qualitative study also identified that a lack of knowledge was a contributor to the slow pace of uptake of automation tools amongst guideline developers and concluded automation tools needed to be transparent and in line with current values and practice.(9)

The interest in systematic review automation is long-standing, and may be dated back to the release of the first version of RevMan in 1993.(14) Its broadening is reflected in the formation of the International Collaboration for the Automation of Systematic Reviews (ICASR) in 2015.(4) Nevertheless, one of the key barriers to the use and adoption of automation tools remains the lack of knowledge about their existence. This is a crucial barrier, as the majority of respondents to our survey self-teach the use of the tools, and they cannot teach themselves to use the tools they do not know exist. This suggests that an increased emphasis on tool dissemination (e.g. such as the SR Toolbox initiative, or through integration into the Cochrane, Campbell, and JBI guidelines and teaching materials) could be prioritised. The 6 organisations we surveyed could also have a role in better dissemination. As one of the other key barriers identified was the steep learning curve, the provision of resources such as demo videos and tutorials for the tools is likewise crucial – in particular as the time investment in acquiring the familiarity with these tools seems to be well offset by the time savings perceived by the respondents.

Automation tools are likely to increase in range and improve in usability, and so take on an increasingly important role in high quality and timely reviews. Regular surveys of their uptake, problems, and abandonment will be important to understanding their dissemination.

## Data Availability

The data collected as part of this study are available on reasonable request from the corresponding author, AMS.

## Acknowledgements

We would like to thank Ton Kujipers for invaluable feedback on the drafts of the survey, and Melanie Vermeulen for setting up the survey in SurveyMonkey. We would also like to thank all of our respondents for generously sharing with us their time, views and suggestions.

## Appendix A The full survey instrument

1. Are your reviews predominantly conducted through an organization such as:
  a. Cochrane
  b. JBI
  c. Campbell
  d. Own Organization
  e. Other
2. Why do you predominantly conduct reviews?
  a. As part of a guideline development process
  b. As part of the Health Technology Assessment (HTA) process
  c. As a systematic review only
3. What types of reviews do you conduct predominantly?
  a. Interventions
  b. Diagnostic Test Accuracy
  c. Prognostic
  d. Etiology
  e. Qualitative
  f. Scoping
  g. Prevalence
  h. Economic Evaluations
  i. Health Utilities
  j. Patient Preferences and Values
  k. Other
4. What stage/s of the systematic review are you most commonly involved in? (select one or more)
  a. Formulating the question (e.g. PICOT)
  b. Search design and/or execution
  c. Screening
  d. Data extraction (including risk of bias)
  e. Data synthesis/meta-analysis
  f. Writeup
  g. Developing recommendations from the evidence
5. How many systematic reviews have you been involved in?
  a. 0
  b. 1-2
  c. 3-5
  d. 6-10
  e. 10+
6. Which tools do you actively use in the systematic review process?
  a. CADIMA
  b. CM-UCLII
  c. Colandr
  d. Covidence
  e. Data Abstraction Assistant (DAA)
  f. Disputatron
  g. Docear
  h. DoctorEvidence (DOC Data)
  i. Engauge Digitizer
  j. EPPI-Reviewer
  k. EXACT: EXtracting Accurate efficacy and safety information from ClinicalTrials.gov
  l. ExaCT: extraction of clinical trial
  m. Fiddle
  n. GRADEpro
  o. Graph2Data
  p. GRIM (granularity-related inconsistency of means) test calculator
  q. Health Assessment Workspace Collaborative (HAWC)
  r. Import.io
  s. JBI-SUMARI
  t. MAGICapp
  u. metaDigitise
  v. ORBIT (Outcome Reporting Bias in Trials) Matrix Generator
  w. Plot Digitizer
  x. Polyglot
  y. ProxyPaper
  z. ReLiS
  aa. Review Manager (RevMan)
  bb. RobotSearch
  cc. RobotReviewer
  dd. ROBVIS
  ee. Scholarcy
  ff. SESRA
  gg. SRDB.PRO
  hh. SWIFT-Review
  ii. SyRF: Systematic Review Facility
  jj. SRA-Helper (Endnote Helper)
  kk. Systematic Review Accelerator
  ll. Voyant Tools
  mm. WebPlotDigitizer
  nn. Weka
  oo. Word Frequency Analyser
  pp. WordStat 8
  qq. Other (please specify)
7. Which three tools would you use most commonly? [text box to fill in]
8. Which tools have you tried to use in the past but discontinued using, and why (optional)? [text box]
9. Of all the systematic reviews (guidelines, HTAs) you have completed in the past 3 years, approximately what percentage involved automation tools:
  a. 0%
  b. 1-25%
  c. 25-50%
  d. 50-75%
  e. 75-100%
10. What stage/s of the systematic review/guideline/HTA were these tools used:
  a. Formulating the PICOT question
  b. Search design and/or execution
  c. Screening
  d. Data extraction (including risk of bias)
  e. Data synthesis/meta-analysis
  f. Writeup
  g. Developing recommendations from the evidence
11. How much time would you say those tools speed up the review process (optional)
  a. Saves lots of time
  b. Saves some time
  c. Neutral
  d. Costs some time
  e. Costs lots of time
12. Do these tools improve the accuracy of the systematic review/guideline/HTA?
  a. A lot of improvement in accuracy
  b. A little improvement in accuracy
  c. Neutral
  d. Lose some accuracy with using the tools
  e. Lose a lot of accuracy with using the tools
13. What prevents you from using the automation tools?
  a. The learning curve (too complicated to learn)
  b. Time constraints
  c. Costs
  d. Language barrier
  e. Internet connectivity/firewall
  f. User unfriendly
  g. Lack of knowledge of existing tools
  h. Other [optional ‘please share why’: text box]
14. What is the primary way in which you learned how to use the automation tools?
  a. Workshops
  b. Webinars
  c. Presentations
  d. Colleague showed me
  e. Help documentation
  f. Self-taught
15. What stage of the review is most time consuming for you?
  a. Formulating the PICOT question
  b. Search design and/or execution
  c. Screening
  d. Data extraction (including risk of bias)
  e. Data synthesis/meta-analysis
  f. Writeup
  g. Developing recommendations from the evidence
16. What automation tools would you like to see in the future – please be specific as possible? [multi-line text box] E.g. “a tool to help with dispute resolution for search”, “a visualisation utility to optimise search strategies”, “an automated method to extract risk of bias”
17. Optional: Please provide your email if you would like to receive the results of the survey (if you would like to remain anonymous but receive the results, please input a free email address such as gmail or hotmail, rather than your institutional address): [text box]

## Appendix B Expanded results

**Table B1:** What types of reviews do you predominantly conduct? Responses under the “other” category, included: network meta-analyses, umbrella reviews, rapid reviews, cost and resource use, health policies, implementation science, investigative tests, mathematical modelling, meta-research, methodological, mixed methods, observational, IPD, risk factor characteristics, systematic maps, to underpin clinical evidence report, and variable.

| Type of review | Responses |  |
| --- | --- | --- |
| Interventions | 82% | 206 |
| Diagnostic test accuracy | 25% | 62 |
| Prognostic | 17% | 44 |
| Etiology | 7% | 18 |
| Qualitative | 29% | 72 |
| Scoping | 46% | 115 |
| Prevalence | 12% | 30 |
| Economic evaluations | 10% | 26 |
| Health utilities | 10% | 24 |
| Patient preferences and values | 11% | 27 |
| Other (please specify) | 10% | 25 |
|  | Total | 252 |

**Figure B1:**
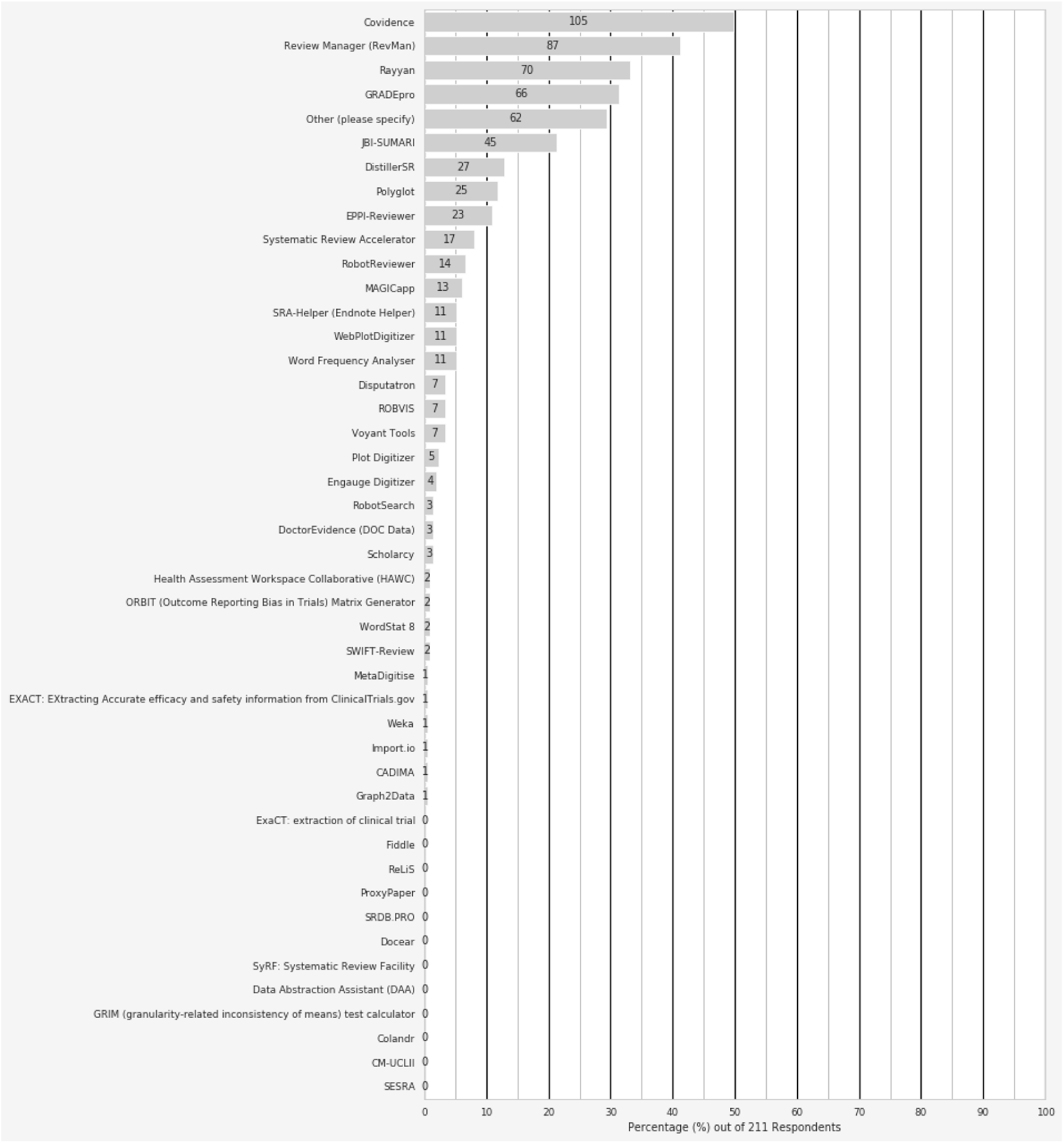
Most commonly used systematic review automation tools – expanded figure. Tools mentioned under the ‘other’ category, included: 2D search, Abstrackr, AtlasTi, VOSViewer, SWIFT-Active Screener, bespoke or ‘home-grown’ tools, Cinema, Cochrane Register of Studies, Endnote (17 responses), Yale Analyser, Vos viewer, PubMed reminer, Yale Mesh, Termine, Cochrane Screen4Me, MeSH on Demand, PubVenn, Epistemonikos, Excel, Zotero, Google translate, Ovid Launcher, Open Access Button, Meta-lite, NVivo, PRISMA, R, Rayyan, RCT Classifier, Revtools, litsearchr, SRDR, STATA, OpenMeta, Sysrev.com, litsearchr, VosViewer, CiteSpace

**Table B2: Tool development suggested by the respondents – expanded table**

**Table 6:** Tool development suggested by the respondents.

| *What automation tools would you like developed?* |  |
| --- | --- |
| **Review stage (N suggestions)** | **Suggested tools\*** |
| Searching (n=53) | • **Strategy optimisation tool (non-specific or visual) (n=14)**<br>• **Deduplication (n=9)**<br>• **Translation between databases (n=7)**<br>• **Strategy design (non-specific or data mining tools) (n=5)**<br>• **Grey literature (non-specific or exporting) (n=2)**<br>• **Synonym/related term (phrase) suggestion tool (n=2)**<br>• Automated study identification (e.g. for living reviews/updates)<br>• For open peer review comments, article versions<br>• For systematic reviews<br>• Formatting abilities (colour coding, notes, comments, indenting)<br>• Full text retriever<br>• Interfaces (better and more stable)<br>• Meta search with filters<br>• OVID interface with federated search function (allowing searches of multiple databases)<br>• Reference importing from reference manager<br>• Searching (non-specific)<br>• Specific features (export results from databases and other sources; ability to group, sort, select results; interoperable with reference management software)<br>• String logic check tool<br>• Subject heading translation between databases<br>• Text mining from relevant papers |
| Screening (n=20) | • **Dispute resolution tool (non-specific or with visualisation) (n=5)**<br>• **Screening tool (non-specific) (n=4)**<br>• **Data mining functionality (for full-text screen) (n=2)**<br>• **Reference prioritisation and threshold-setting functionality (n=2)**<br>• Automated ti-ab scan for keywords/exclusion criteria<br>• Data mining functionality (for full-text screen)<br>• Decision tracking functionality (inclusion/exclusion decisions)<br>• Filtering out RCTs<br>• Guided algorithm tool<br>• Pdf importer from citation managers<br>• Search result clustering functionality<br>• Study selection tool (non-specific) |
| Data extraction (other than risk of bias) (n=41) | • **Data extraction tool (non-specific or general - e.g., accurate, fast) (n=24)**<br>• **For observational studies (n=2)**<br>• **For reuse of previous data extractions (n=2)**<br>• **From Pdfs (n=2)**<br>• Customisable<br>• Dispute resolution tool for data-extraction<br>• Double blind extraction capability<br>• For descriptive data<br>• For effect sizes |
|  | • For meta-data<br>• For outcome data<br>• From figures/tables<br>• Identifies relevant parts of the article<br>• Of references within systematic reviews for import into reference manager<br>• Text mining capability |
| Risk of Bias (n=19) | • **Automated risk of bias (non-specific) (n=11)**<br>• **Automated risk of bias (observational studies) (n=7)**<br>• Automated strength of evidence assessment |
| Synthesis/analysis (n=5) | • Non-specific<br>• Importing into a manuscript (like RevMan exporter)<br>• Better capacity in DistillerSR<br>• Topic modelling<br>• Results visualisation |
| Writeup (n=12) | • **Data visualisation/presentation (n=5)**<br>• Automated reported checklist completion<br>• Correctness check (Cis and effect sizes)<br>• Figure generation<br>• PRISMA generator<br>• Table generator<br>• Write-once-read-many functionality<br>• Writing from standardised template |
| Project management (n=16) | • **Streamlined/single tool for the entire process (n=9)**<br>• **Linkage between tools/data pushing between tools (n=2)**<br>• Automated pre-registration forms<br>• Collaborative work tool (facilitate work, email tracking)<br>• Paper trail tracking<br>• Reference management tool<br>• Tools (non-specified) for qualitative evidence reviews |
| Other (n=4) | • Causation assessment<br>• Extracting meta-data (checklist information)<br>• Free storage (for articles)<br>• Unclear (literature selection tools) |

\*where more than one suggestion mentioned a specific tool, the number of suggestions is indicated in brackets; most common suggestions are listed first and bolded

